# Social concepts rely on a domain-general anterior-temporal hub and social “spokes” in ventral prefrontal cortex and insula

**DOI:** 10.64898/2026.07.02.26357102

**Authors:** Matthew A. Rouse, Peter Garrard, James B. Rowe, Matthew A. Lambon Ralph, Timothy T. Rogers

## Abstract

A long-standing debate surrounding the neural bases of social concepts concerns the role of anterior temporal lobe (ATL). One perspective suggests ATL subregions are dedicated specifically to social knowledge; another suggests the ATLs constitute a domain-general hub for conceptual knowledge, but with graded functional specialisation depending on connectivity to modality specific “spokes.” The positions have been difficult to adjudicate due to many confounding factors in tests of social and non-social knowledge. We address these challenges via three innovations in assessment of knowledge in frontotemporal dementia (FTD). First, we introduce a new task that controls for several potential confounds. Second, we apply mixed linear models to behavioural data analysis, allowing further control over confounding factors. Third, we extend the mixed-model approach to lesion-symptom mapping, identifying cortical regions where structural pathology yields a disproportionate impairment on social versus non-social knowledge when other factors are controlled. We used these techniques to probe social and non-social knowledge in FTD subtypes: semantic dementia (SD), associated with asymmetric-bilateral ATL atrophy (n=21), and behavioural-variant (bvFTD), characterised by frontoinsular atrophy (n=24). When confounding factors were controlled, people with SD showed an equal impairment for social and non-social concepts, whereas those with bvFTD were disproportionately impaired on social concepts. The *differential* impairment of social concepts was associated with atrophy in the insula, orbitofrontal and ventromedial prefrontal cortex and other regions implicated in social knowledge generally. The results suggest that the bilateral ATLs constitute a domain-general semantic hub, whereas ventral prefrontal and insula cortex contribute preferentially to knowledge about people.

**Open Access:** For the purpose of open access, the UKRI-funded authors have applied a CC BY public copyright licence to any Author Accepted Manuscript version arising from this submission.

## 1. Introduction

A persistent debate in theories about the neural bases of human conceptual knowledge concerns the role of the anterior temporal lobes (ATLs), where two somewhat different views have emerged across literatures. In social cognitive neuroscience, the *social knowledge hypothesis* suggests that the ATLs may be divided into distinct, functionally-specific subregions, some of which are specialised to represent social concepts; that is, knowledge relevant to our understanding of/interactions with other people (Olson et al., 2013; Ross & Olson, 2010; Simmons & Martin, 2009; Zahn et al., 2007, 2009). Work in semantic representation more broadly, however, argues that the ATLs constitute a *domain-general hub* that supports knowledge for all conceptual domains across all modalities of reception and expression (Patterson et al., 2007), showing only graded functional specialisation that reflects its connectivity to other representational modalities, e.g. vision, language, action, etc. (Binney et al., 2012; Lambon Ralph et al., 2017; Rice, Hoffman, et al., 2015; Visser & Lambon Ralph, 2011). On this view the ATL’s contribution to social-semantic processing reflects its broad role in semantic cognition generally, rather than the participation of modular sub-regions in a domain-specific social-knowledge network (Binney & Ramsey, 2020; Rouse, Binney, et al., 2024).

Both views cite empirical support. Regarding the social knowledge hypothesis, many studies report ATL subregions where functional activation correlates with a variety of social-semantic tasks including theory-of-mind (Balgova et al., 2022, 2024; Frith & Frith, 2003), processing of social word-meanings (Zahn et al., 2007), recognition and retrieval of knowledge about individual people from face images or names (Collins & Olson, 2014), perception of Heider and Simmel animations depicting abstract social interactions (Ross & Olson, 2010), and moral judgments (Moll et al., 2005). Patients with anterior temporal pathology arising from frontotemporal dementia (FTD) show deficits recognising and retrieving information about people from face images and from names (Borghesani et al., 2019; Ding et al., 2020; Rouse, Ramanan, et al., 2024; Snowden et al., 2004), as do patients following anterior temporal resection to remediate epilepsy (Drane et al., 2013; Glosser et al., 2003; Rice, Caswell, et al., 2018; Rouse, Ramanan, et al., 2024). Some lesion-symptom correlational results suggest that impairment to social concepts correlates reliably with hypometabolism in the right superior ATL, a result that directly mirrors patterns of functional activation evoked by processing the meanings of social words in at least some fMRI studies (Zahn et al., 2007).

Regarding the semantic-hub hypothesis, a host of functional brain imaging studies show increased activation in the ventral ATLs either bilaterally (Binney et al., 2010; Visser, Embleton, et al., 2010) or in the left (Ross & Olson, 2010) for semantic relative to non-semantic processing of words and pictures. This applies for a wide range of semantic domains, and for abstract, concrete, and social items (Rice, Hoffman, et al., 2018) so long as the imaging methodology takes measures to resolve signal in this area (Halai et al., 2014; Visser, Jefferies, et al., 2010). Transcranial magnetic stimulation (TMS) applied to both the left and right ATLs produces significant slowing of semantic decisions for both word and image comprehension, and for both abstract and concrete semantic domains including social concepts (Pobric et al., 2009, 2010, 2016). Anterior temporal pathology in FTD erodes knowledge across all receptive and expressive modalities, and for all tested conceptual domains including social concepts (Rijpma et al., 2023; Rouse, Halai, et al., 2024). Moreover, the resulting deficits show a relative sparing of highly familiar and/or prototypic concepts, and of very broad semantic distinctions, with a profound impairment to knowledge about specific, unfamiliar, and atypical semantic categories (Rogers et al., 2015); and similar, albeit more subtle deficits are also observed following anterior temporal resection (Lambon Ralph et al., 2012; Rice, Caswell, et al., 2018; Rouse, Halai, et al., 2024; Rouse, Ramanan, et al., 2024).

Adjudicating the two positions via functional brain imaging and/or comparative patient studies has proven more challenging than it might at first appear. Human semantic processing can be influenced by myriad cognitive and psycholinguistic factors that are difficult to control across social and non-social stimuli, including abstractness, frequency, contextual diversity, and executive factors (see e.g., Binney et al. 2016). Tasks used to assess knowledge of social versus non-social concepts often differ in many respects, potentially placing differential demands on strategic, attentional, perceptual, affective, or linguistic processes. Also of key importance, definitions of what “counts” as a social concept can be somewhat loose and variable across studies (Pexman et al., 2023; Rouse, Binney, et al., 2024; Rouse, Halai, et al., 2024). Thus, where patient or neuroimaging data appear to show different patterns of activation or dysfunction for social versus non-social items, it has been difficult to rule out the influence of many confounding factors.

Perhaps for these reasons, the few studies directly contrasting social and non-social knowledge have painted a mixed picture. The early work from Zahn and colleagues (2007) cited above reported greater activation for social compared to non-social concepts in the right superior ATL, with subsequent work in participants with FTD showing that hypometabolism in this region correlates more with social than non-social knowledge impairments (Zahn et al., 2009). Yet subsequent studies, arguably using better-controlled materials, found that the right-superior imaging result replicates only for highly abstract social concepts (Rice, Hoffman, et al., 2018); or that the social-semantic peak arises in the left hemisphere (Ross & Olson, 2010); or that it appears both bilaterally and in superior and inferior aspects (Binney et al., 2016), raising questions about both the nature and the location of the effect. While anterior temporal regions undoubtedly contribute to recognition/retrieval of information about familiar people, both imaging and patient studies suggest the same regions also contribute to recognition/retrieval about any familiar unique entity (Rogers et al., 2015), and indeed about more general concepts, showing gradedly stronger responses when participants must categorise stimuli with high precision (e.g. as a “magpie” rather than as a “bird” or an “animal”) (Rogers et al., 2006). Thus, it is unclear whether the activation and deficit patterns reflect a special role for the ATLs in social processing, or increased demands on a domain-general hub as tasks require greater semantic precision (Rogers et al., 2015). Most recently, a large-scale study assessing people with FTD and with anterior temporal resection on a broad array of social, semantic, and other cognitive tasks found no difference between social and non-social task performance when other aspects of the neuropsychological profile were accounted for, consistent with the view that social and non-social knowledge rely on the same neurocognitive systems (Ding et al., 2020; Rice, Caswell, et al., 2018; Rouse, Halai, et al., 2024).

*Both* proposals are at odds with common FTD presentations at clinic, which, while often mixed, can vary between two quite different forms. Patients with *semantic-variant* primary progressive aphasia (svPPA), often known as *semantic dementia* (henceforth SD), typically present with word-finding complaints. On subsequent testing such patients display a broader semantic impairment encompassing multiple modalities and conceptual domains that is accompanied by grey matter thinning and/or hypometabolism focused on the anterior temporal cortices bilaterally (Gorno-Tempini et al., 2011; Mummery et al., 2000). While face/person recognition is frequently impaired early on (Ding et al., 2020; Evans et al., 1995; Gorno-Tempini et al., 2004), social dysfunction is not a characteristic presenting symptom for such patients. When the semantic dementia has right-predominant atrophy (right-SD, also known as right temporal variant FTD, rtvFTD), early changes in social behaviour are more common and may lead to misdiagnosis as the behavioural-variant FTD.

The *behavioural-variant* of FTD (henceforth bvFTD) typically presents with primary complaints of socially inappropriate behaviours and personality change (Rascovsky et al., 2011). On subsequent testing such patients show characteristics of executive dysfunction accompanied by grey matter thinning and/or hypometabolism consistently focused in inferior frontal regions (including orbitofrontal/ventromedial prefrontal cortex and anterior insula) (Ilchovska et al., 2026). While the presenting behavioural symptomatology is often attributed to the disinhibition that characterises prefrontal disorders generally (Hughes et al., 2015), it might equally or alternatively arise because these regions contribute preferentially to social conceptual knowledge. Indeed, this possibility accords with a broad literature implicating the orbitofrontal cortex in aspects of motivation, reward monitoring, and affect - all varieties of cognitive functioning that are especially important to social behaviours (Rankin, 2020; Rouse, Binney, et al., 2024). Thus, the literature suggests three distinct hypotheses about the neural bases of social and non-social conceptual knowledge that, in principle, can be empirically adjudicated by comparison of patients with bvFTD versus SD - so long as the assessments permit control over potential confounding factors. Specifically:

1. *ATLs contain a social-knowledge system.* On this view there is a dedicated system for social concepts situated somewhere within the ATLs, possibly in their right superior aspect. Thus when confounding factors are controlled (a) patients diagnosed with SD should show worse social-semantic knowledge than those with bvFTD due to increased likelihood of ATL pathology, (b) regardless of diagnosis, patients with predominantly temporal lobe pathology should show worse social knowledge than those with predominantly frontal pathology, and (c) lesion-symptom mapping should identify some ATL subregions, including right superior aspects, as differentially important for social relative to non-social concepts.
2. *ATLs constitute a domain-general hub.* On this view ATLs support knowledge for all conceptual domains equally, with no sub-region dedicated to social knowledge specifically. Thus when confounding factors are controlled, (a) patients diagnosed with SD should show equally poor knowledge of social and non-social domains, with both worse compared to those diagnosed with bvFTD, (b) regardless of diagnosis, patients with predominantly temporal pathology should be more impaired than those with predominantly frontal pathology on both social and non-social semantic tasks, and (c) lesion-symptom mapping should not identify any ATL subregions as differentially important for social relative to non-social concepts.
3. *ATL-hub plus inferior frontal/insula “spokes” for social knowledge.* On this view ATLs are equally important for social and non-social concepts while orbitofrontal, ventral prefrontal and insula cortices contribute more to social than to non-social knowledge by virtue of their role in motivation, reward, and affect processing. Thus when confounding factors are controlled (a) patients diagnosed with SD should show an equal impairment for social and non-social domains while those diagnosed with bvFTD should show worse performance on social than non-social items, (b) patients with predominant ATL pathology should show equal impairment of social and non-social items while those with frontal-predominant pathology should show worse impairment of social items and (c) lesion-symptom mapping should identify inferior/ventral prefrontal and insula regions as differentially important for social relative to non-social concepts.

The current paper aims to adjudicate these hypotheses via three innovations in the assessment of social and non-social knowledge in patients with FTD that remediate the challenges noted earlier. First, we introduce a new word-to-picture matching assessment that (a) focuses on *well-defined* social and non-social subdomains, and (b) matches social and non-social items for several factors known to influence behaviour in semantic matching tasks. Second, after testing a large cohort of bvFTD and SD patients on the new assessment and many other measures, we apply a series of *nested binomial mixed-effects models* that allow us to regress out the effects of neuropsychological and item-wise cognitive factors known to influence semantic performance. This provides a principled means of then evaluating whether the factors of theoretical interest - the patient diagnosis (SD vs. bvFTD) and the stimulus type (social/non-social) help to predict performance after other explanatory factors have been accounted for. Third, we introduce a novel *mixed-linear lesion-symptom mapping* technique that allows us to identify regions of cortex where pathology has a disproportionate impact on social versus non-social test items *after* the effects of other factors have been regressed out - ensuring that the resulting maps do not reflect correlations between grey matter pathology and other potentially confounding factors. Together the results suggest a view of conceptual representation in the brain that reconciles literature in social and semantic cognition.

## 2. Methods

The *social-roles* assessment evaluates knowledge about the meanings of nouns referring either to classes of people or classes of manmade objects, using a four-alternative forced-choice word to picture matching task. On each trial, the word appears above four photographs and the respondent must choose which image best matches the word. In the *social* condition, each word denotes a class of person with an implied age (e.g. “adult,” “toddler,” “pensioner”) or both an implied age and gender identity (e.g. “lad,” “debutante,” “widow”). All photographs show images of people, with distractors selected to neighbour the target in age and gender. For example, the target for the word “grandma” shows an older woman, while the distractors included a young woman (gender match), an older man (age match), and a young man (neither matching). For the gender-neutral word “toddler”, the target shows a boy about age 2 and the distractors include an infant (near-age, younger), an older boy (near-age, older), and an adolescent boy (farther-age). Thus, successful performance requires, not just knowing that a word refers to a person but understanding it’s meaning well enough to discriminate the target from close semantic associates. In the *non-social* condition, each word names a kind of manmade object, for instance, *screwdriver, shed, cup* etc - with distractors selected from the same intermediate category. Thus for “screwdriver,” distractors are all handheld tools: a hammer, saw, and spanner; for “shed” they are all enclosures: house, church, tent; etc. Successful performance again requires sufficient understanding to discriminate the target from close semantic neighbours.

The task is unambiguously semantic in requiring comprehension of a word meaning and cross-modal matching to an image; indeed, word-picture matching is perhaps the most common tool for assessing disorders of semantic memory. Words referring to people are, by definition, more “social” than words referring to inanimate objects, so the distinction between social and non-social in this task context is clear. The social features implied by the target words - age and perceived gender identity - are deeply engrained properties of person-knowledge acquired early in development (Shutts et al., 2010) and govern the similarities participants discern amongst people (Colón et al., 2024). Both social and non-social conditions require comprehending a single word and matching it to one of four photographs, all depicting items from the same semantic category. Moreover, each target category appears equally often as a distractor across other items; for instance, a picture of a key appears as the target for the “key” item, but three other pictures of different keys appear as distractors for three other items, so correct choices cannot be inferred by the frequency or repetition of a given category, and no image is repeated. In this sense, strategic, attentional, perceptual and other cognitive task demands are matched.

Furthermore, all items require only single-word comprehension, and all words are of the same grammatical class, minimising reliance on syntactic or other aspects of linguistic processing. Both tasks require discrimination of the target from near semantic neighbours, i.e., high semantic precision—a factor that strongly influences performance in all varieties of semantic impairment but often goes uncontrolled (Rogers et al., 2015). Finally, words in each condition were matched for log frequency and, critically, accompanied by measures of other important confounding factors known to impact comprehension: concreteness and contextual variability (i.e., the degree to which a word’s meaning depends on its context; see Hoffman et al. 2013). Thus, the task permits analysis via mixed linear models that allow us to regress out possible confounds both by patient and, importantly, by-item, greatly improving power to detect any theoretically important effects of diagnosis (SD vs. bvFTD), condition (social/non-social), or their interaction.

The logic of the study is as follows. We assessed a large cohort of patients diagnosed with FTD on the social-roles task and a battery of other semantic and cognitive measures previously described by Rouse et al. (2024), as well as collecting structural brain scans for a subset of participants. In a behavioural analysis, we first fitted a *baseline* logistic mixed effects model to predict a participant’s accuracy on every individual test item (a binary outcome) from neuropsychological and stimulus factors known to impact performance on semantic tasks: the overall magnitude of the semantic impairment (assessed across a variety of verbal and non-verbal tasks but excluding the social-roles assessment), and word frequency, concreteness and semantic diversity. We then assessed whether the model fit improved significantly when diagnosis (SD/bvFTD), stimulus type (social/non-social), and their interaction were added.

Note that this comparison adjudicates the three hypotheses listed earlier. The domain-general hub hypothesis stipulates that, once the effects of stimulus factors like frequency, concreteness, and semantic diversity are accounted for, accuracy on both social and non-social items should be determined solely by the overall magnitude of the domain-general semantic impairment. Thus, adding diagnostic category, stimulus type, and their interaction should *not* improve model fit. In contrast, the social knowledge hypothesis predicts that, after these effects have all been regressed out, performance on social items will still additionally depend on the degree to which the social-knowledge subsystem within the ATLs has been impaired. Since ATL pathology is a diagnostic feature of SD, this predicts that the interaction of diagnosis and stimulus type should explain additional variance in task performance, with SD patients showing comparatively *worse* performance on social than non-social items. Under the OFC-spoke hypothesis, knowledge for all domains depends upon the ATL, but social concepts depend comparatively more on the inferior/ventral PFC and insula - thus, after overall semantic impairment and other factors have been regressed out, the interaction of diagnosis and stimulus type should also account for significant additional variance, but in the reverse direction, with bvFTD patients showing comparatively worse performance on social than non-social items. Following this analysis, we additionally considered whether the resulting effects persisted when other aspects of the neuropsychological profile, including executive dysfunction and overall cognitive decline, were added to the model.

Following the behavioural analysis, we considered how patterns of dysfunction relate to underlying structural pathology across patients. First, we used standard techniques to compare the patterns of atrophy associated with each diagnostic category, as well as patterns associated with each aspect of the neuropsychological profiles (semantic impairment, executive impairment, and overall cognitive decline). We then introduce a novel analysis that extends the mixed-linear model to lesion-symptom mapping, allowing us to visualise where cortical neuropathology predicts differential impact on social versus non-social items in the social-roles assessment.

### 2.1. Participants

We recruited forty-five people with a clinical diagnosis of probable frontotemporal dementia (semantic dementia = 21, bvFTD = 24) from specialist clinics in Addenbrooke’s Hospital, Cambridge (*n* = 37), St George’s Hospital, London (*n* = 4) and John Radcliffe Hospital, Oxford (*n* = 4). Nineteen age-matched healthy participants were recruited from the volunteer panel at the MRC Cognition and Brain Sciences Unit, University of Cambridge. Most participants provided informed consent obtained according to the Declaration of Helsinki. If participants lacked capacity to consent, their next of kin was consulted using the ‘personal consultee’ process as established by UK law. Demographic and disease information is displayed in Table 1.

**Table 1.** Demographic and disease information.

|  | SD | bvFTD | Control | Group difference | Post-hoc difference |
| --- | --- | --- | --- | --- | --- |
| N | 21 | 24 | 19 | - | - |
| Sex (M:F) | 8:13 | 16:8 | 9:10 | $\chi^2 = 3.85$, $p = 0.15^a$ | - |
| Age (years) | 66.0 (6.9) | 64.8 (9.2) | 64.4 (6.7) | $F(2,61) = 0.22$, $p = 0.80^b$ | - |
| Education (years) | 13.5 (2.9) | 11.3 (1.6) | 15.6 (3.4) | **$F(2,60) = 13.62$, $p$** | bvFTD, SD < C; bvFTD < SD |
| Years since symptom onset | 5.7 (3.3) | 5.9 (3.6) | - | $W = 236.5$, $p = 0.92^c$ | - |
| Years since diagnosis | 2.2 (1.6) | 2.6 (1.6) | - | $W = 311$, $p = 0.18^c$ | - |

<sup>a</sup>Chi-square test, <sup>b</sup>one-way ANOVA, <sup>c</sup>Wilcoxon rank-sum test
Mean and standard deviations are reported for each group. Significant p-values are highlighted in bold.
bvFTD = behavioural-variant frontotemporal dementia, C = control, SD = semantic dementia

### 2.2. Structural MRI

#### 2.2.1. Acquisition and pre-processing

Thirty-four FTD participants (SD = 19, bvFTD = 15) and 35 controls had a T1-weighted 3T structural MRI scan on a Siemens PRISMA at the University of Cambridge, using an MPRAGE sequence. Thirty participants (FTD = 14, control = 16) were scanned at the MRC Cognition and Brain Sciences Unit (repetition time (TR) = 2000ms, echo time (TE) = 2.85ms, inversion time (TI) = 850ms), whereas 39 participants (FTD = 20, control = 19) were scanned at the Wolfson Brain Imaging Centre (TR = 2000ms, TE = 2.93ms, TI = 850ms). Raw data were converted to the Brain Imaging Dataset format and preprocessed using the Computational Anatomy Toolbox version 12 in SPM12. Images were segmented into grey matter, white matter and CSF, modulated, and normalised to MNI space using geodesic shooting. Normalised grey matter images were spatially smoothed using a 10mm full width half maximum Gaussian kernel.

#### 2.2.2. Grey matter volume differences between groups

Voxel-based morphometry was used to explore differences in grey matter volume between each FTD group and controls. Separate general linear models were fitted for each contrast, with age, intracranial volume and scanner site included as covariates. Independent t-tests were conducted between (i) SD vs. controls and (ii) bvFTD vs. controls. An explicit mask was used, based on a method recommended for severely atrophic brains (Ridgway et al., 2009). Significant clusters were extracted using a cluster-level threshold of *q* < 0.05, based on an initial voxel-level threshold of *p* < 0.001. All neuroimaging results were visualised using MRIcroGL and AFNI Surface Mapper (Saad et al., 2004), and brain regions were labelled using the Automated Anatomical Labelling Atlas.

The grey matter differences between each FTD group and controls are displayed in Fig. 1 and reported in Supplementary Table 1. Each FTD group had maximal volume loss in their associated atrophy centres (i.e., bilateral ATL atrophy in SD, and frontoinsular atrophy in bvFTD). However, there was overlapping atrophy between the two groups, with prefrontal cortical volume loss in SD, and bilateral ATL atrophy in bvFTD. This finding highlights the limitations of diagnosis-based group comparisons for disentangling the precise contributions of different brain regions to social conceptual knowledge in FTD and gives justification for the use of a transdiagnostic approach (Ding et al., 2020; Murley et al., 2020; Ramanan et al., 2023; Rouse, Halai, et al., 2024).

**Fig. 1.**
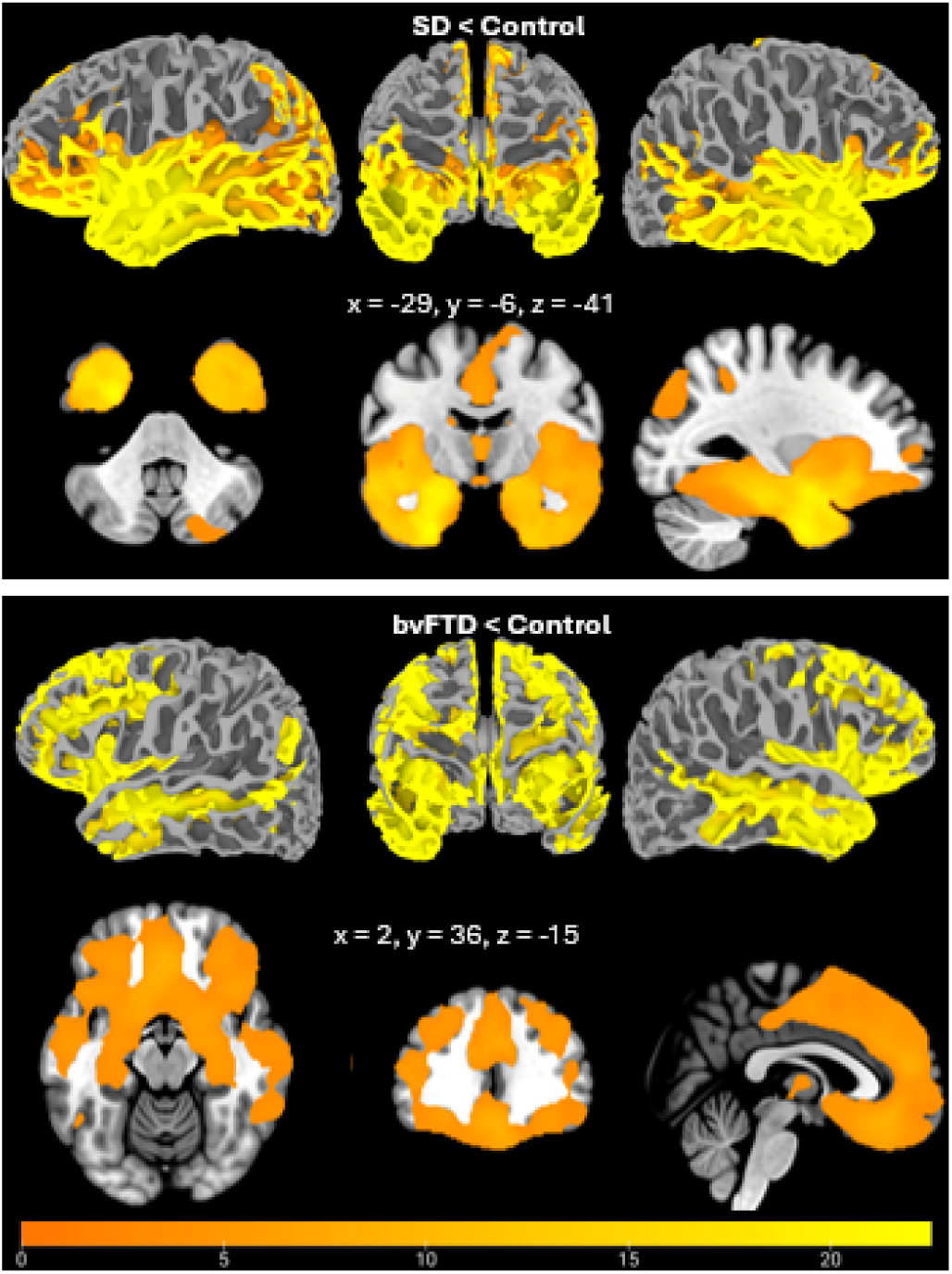
Regions of significantly reduced grey matter intensity in SD (top) and bvFTD (bottom). In each panel the top row shows regions of significant atrophy projected onto a standard surface model, while the bottom row shows slices through a standard brain volume at the coordinates indicated. While the SD group shows more temporal and less frontal involvement than the bvFTD group, both show significant atrophy in both anterior temporal and prefrontal regions.

### 2.3. Neuropsychology

#### 2.3.1. Background neuropsychology

Participants completed a detailed test battery, including comprehension of multiple types of social concepts, including famous people, abstract social concepts, emotion knowledge, social norms understanding and sarcasm detection. Where possible, ‘non-social’ comparator tasks were included, matched for psycholinguistic variables such as frequency, imageability and specificity. General semantic memory was assessed using the modified picture version of the Camel and Cactus Test (CCT), and naming task from the Cambridge Semantic Memory Test Battery. Executive function was assessed using the Brixton Spatial Anticipation Test and Raven’s Coloured Progressive Matrices Set B (Raven et al., 1962). Global cognition was assessed using the Addenbrooke’s Cognitive Examination-Revised (ACE-R), a dementia screening tool with five subscales: Attention and Orientation, Memory, Language, Fluency and Visuospatial Function (Mioshi et al., 2006). For a detailed description of each task, see Rouse et al. (2024).

#### 2.3.2. The social-roles task

We developed a novel four-alternative forced choice 35-item word-picture matching task to assess social and non-social conceptual knowledge in FTD. In each *social* trial, participants see and hear a target word denoting a class of person with a characteristic age and/or gender (e.g., *infant*, *woman*, *uncle*). The response options are four coloured photographs of people neighbouring the target in age and gender. Participants are asked to point to which of the four images best matches the target word. The location of the target image was counter-balanced across trials. Non-social trials used the same procedure but with names and images of inanimate man-made objects (e.g., *screwdriver, shed*). Non-social distractors were drawn from the same semantic category as the target (e.g. tools for screwdriver; enclosures for shed; etc). The tasks were carefully matched for log word frequency, a psycholinguistic factor known to have a strong influence on performance (Rogers et al., 2015). The log frequency of each item in each task was obtained from the British National Corpus, and each social item was paired with a non-social item with a similar frequency. Each target word was also associated with a concreteness rating obtained from Brysbaert et al. (2014), allowing us to regress out effects of this by-item factor in the mixed-effects model (Brysbaert et al., 2014).

### 2.4. Statistical analysis

#### 2.4.1. Background neuropsychology

Neuropsychological group comparisons were conducted using the ‘rstatix’ package in R studio version 4.0.3. Normality of data and equality of variance were assessed using Shapiro-Wilk tests and Q-Q plots. Where data were normally distributed, one-way ANOVAs and post-hoc Tukey’s range tests were conducted. In cases where data were not normally distributed, Kruskal-Wallis tests and post hoc Dunn’s tests were conducted. For all tests, a threshold of *p* < 0.05 was used to determine statistical significance.

A varimax-rotated principal component analysis was conducted on the FTD background neuropsychological scores to extract the cognitive dimensions explaining the variance in the data, following the procedure reported by Rouse et al. (2024) with the same data but excluding the social-roles test (Rouse, Halai, et al., 2024). Briefly, the number of principal components was determined using the elbow method on the scree plot of eigenvalues. Factor scores were calculated using the regression method, and sampling adequacy and suitability of the data for PCA were assessed using the Kaiser-Meyer-Olkin test and Bartlett’s test of sphericity. The PCA extracted three principal components, interpreted via the task loadings to reflect composite indices of: (1) *semantic functioning* (including both social and non-social semantic tasks but not the social-roles test), (2) *global cognitive functioning* and (3) *executive functioning.* In all cases higher scores indicate better performance. Scores on these three indices were included as predictors in the linear mixed effects models described in the next section. A linear regression model was fitted to explore the association between grey matter intensity and factor scores on each component, with age, ICV and scanner site included as covariates. Significant clusters were extracted using a cluster-level threshold of *q* < 0.05, based on an initial voxel-level threshold of *p* < 0.001.

#### 2.4.2 Social vs. non-social word-picture matching

A mixed ANOVA was conducted to compare performance between the three groups (SD, bvFTD, controls) on the social vs. non-social word-picture matching tasks. We then fitted a series of hierarchical binomial linear mixed-effects models to predict accuracy for an individual item, across all items (*n* = 70) and the full FTD cohort (*n* = 45). We first developed a *baseline* model assessing contributions from three factors shown to predict semantic performance in prior work: (1) log word *frequency*, which typically predicts higher accuracy under semantic impairment in FTD (Jefferies & Lambon Ralph, 2006; Lambon Ralph et al., 1998); (2) word *concreteness*, which also typically predicts higher performance under semantic impairment in FTD (Hoffman, Jones, et al., 2013; Jefferies et al., 2009); and (3) *semantic functioning* measured as the factor score on the first principal component described above, which self-evidently predicts better performance on semantic tasks like the social-roles test. Following development of the baseline model, we additionally considered a fourth by-item factor: *semantic diversity*, which measures how much a word’s meaning varies across contexts (Hoffman, Lambon Ralph, et al., 2013).

To understand whether these factors together are sufficient to explain performance on the social-roles task (domain-general hub hypothesis) or whether different diagnostic groups show different patterns across social and non-social items (social-knowledge and OFC-spoke views), we next assessed whether baseline model fit improved significantly when *group* (bvFTD, SD), *test-type* (social, non-social), and their interaction were added to the model. *Group* and *test-type* were dummy coded, with SD set as the reference *group*, and ‘non-social’ set as the reference *test-type*.

## 3. Results

### 3.1. Background Neuropsychology

Scores on the background neuropsychology tests are reported in Supplementary Table 2. The SD participants had lower *semantic functioning* scores than bvFTD (*t*(39.1) = 5.57, *p* < 0.0001, *d* = 1.64), indicating worse semantic performance in SD. The bvFTD group had lower scores on the *executive functioning* index (*t*(43) = 3.58, *p* = 0.0009, *d* = 1.06), in line with increased levels of executive impairment in bvFTD. There was no significant difference in factor scores between groups on the *global cognitive functioning* index (*t*(43) = 0.92, *p* = 0.36, *d* = 0.27), indicating that overall cognitive ability was equally compromised in the two groups. Fig. 2 displays the regions of grey matter intensity correlated with performance on the principal components. *Semantic functioning* scores were associated with grey matter volume in the bilateral ATLs, maximal at the temporal pole and ventral ATL and with no frontal involvement. *Global cognitive functioning* scores were correlated with grey matter volume in the precentral gyrus, frontal/orbital gyri, cingulate cortex, insula and supplementary motor area. No clusters were significant for the *executive functioning* factor. Full details of the atrophy correlations are reported in Supplementary Table 3.

**Fig. 2.**
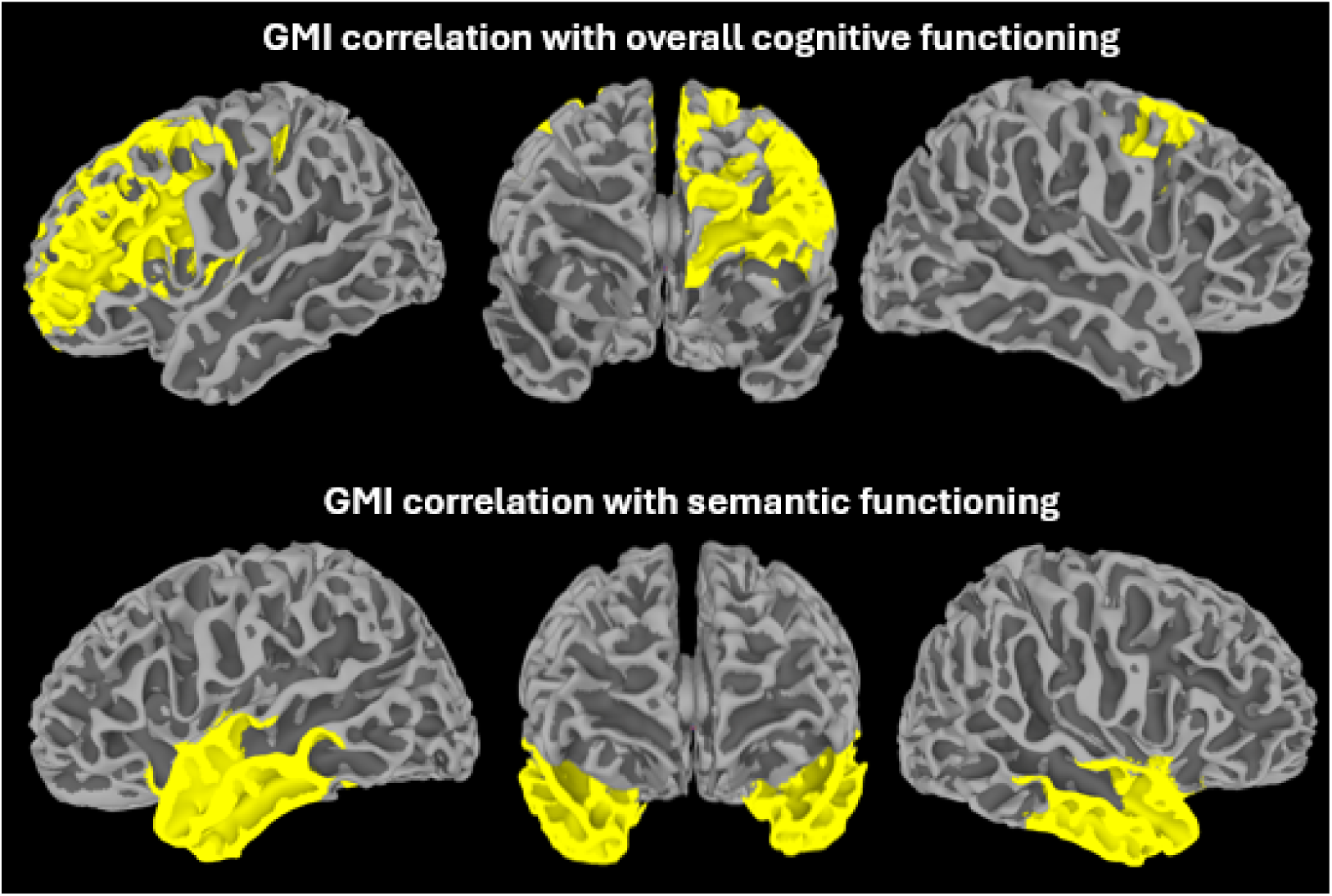
Correlation between grey matter intensity (GMI) and two principal components derived from the cognitive battery: overall cognitive functioning (top) and semantic functioning (bottom). Coloured areas show regions where the correlation between atrophy and metric is significant across participants. Whereas overall cognitive functioning was correlated with pathology in left superior and lateral prefrontal and right superior prefrontal cortex, semantic functioning correlated only with pathology in the anterior temporal lobes bilaterally.

### 3.2. Social concepts word-picture matching

#### 3.2.1. Group comparisons

A mixed ANOVA found a significant group by test-type interaction (*F*(2,61) = 3.98, *p* < 0.009). Post-hoc tests revealed that SD were impaired on both tasks relative to controls (*social*; *p* < 0.0001, *non-social*; *p* = 0.0008), in line with a parallel degradation for both types of semantic knowledge. Relative to controls, bvFTD participants as a group showed significantly worse performance on the *social* task (*p* = 0.007) but not on the *non-social* task (*p* = 0.14). Of course, this result does not accommodate the various confounding factors that the mixed-linear approach is intended to remediate.

#### 3.2.2. Binomial linear mixed effects models

##### Baseline model

All mixed-linear models included random-effect intercepts for item and participants. We first fitted baseline models considering simple effects of each primary factor: semantic functioning factor score (by patient), word frequency and word concreteness (both by item). *Semantic functioning* (beta = 1.14, *z* = 5.49, *p* < 0.0001) and *word frequency* (beta = 0.29, *z* = 3.47, *p* = 0.0005) both accounted for significant variation in participant accuracy, with accuracy log-odds increasing as with higher semantic scores (OR = 3.13) and higher frequencies (OR = 1.34). There was no significant effect of *concreteness* either alone (beta = 0.41, *z* = 1.47, *p* = 0.14) or when added as a simple effect following the other two factors (χ^2^(1) = 2.10, *p* = 0.15). To test whether prediction accuracy improved significantly with the addition of the pairwise interaction term between any of the three fixed factors, we fitted three additional models, each adding a pairwise interaction, and compared its fit to the simple-effects model. None of the interaction terms significantly improved model fit. *Concreteness* was removed, resulting in the following baseline model:

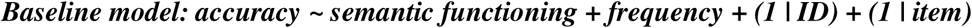

We next considered whether an additional by-item confound might account for additional variation, namely *contextual diversity* - a metric that indicates how much a word’s meaning varies across contexts (Hoffman, Lambon Ralph, et al., 2013). Words with higher diversity are thought to place higher demands on systems of semantic control. Prior work with SD found no impact of contextual diversity on semantic task performance (Hoffman, Lambon Ralph, et al., 2013), and indeed the measure is strongly correlated with concreteness. Yet semantic control does predict performance in patients with acquired deficits of semantic control following stroke (Jefferies & Lambon Ralph, 2006); and social and non-social items may place differing demands on semantic control. We therefore fitted models adding semantic diversity to the baseline model as a by-item fixed effect, either on its own or also including pairwise interactions with frequency and semantic memory measures. In no case did this addition significantly improve model fit, therefore contextual diversity was omitted, and the same baseline model was retained.

### The effect of group and test-type on accuracy

The baseline model accounts for effects of overall semantic memory and word frequency on by-item accuracy in the social-roles task and further establishes that neither concreteness nor contextual diversity predict additional variation on these items. The central question is whether these factors are sufficient to explain performance across both FTD variants for both social and non-social items (per the domain-general hub hypothesis), or whether model fit is improved when diagnosis, stimulus type and their interaction is added (per both social-knowledge and OFC-spoke hypotheses). If these terms do improve model fit, an additional question concerns the direction of the interaction: the social-knowledge hypothesis predicts worse performance on social than non-social items within the SD group, whereas the OFC-spoke hypothesis makes the opposite prediction. We therefore added these terms to the baseline model and evaluated whether they significantly improve model fit:

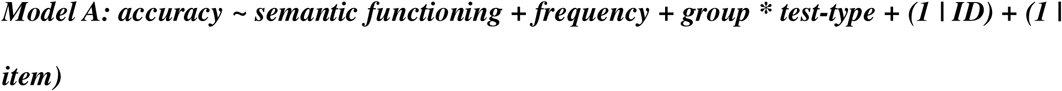

Model A had a significantly increased likelihood than the baseline model (χ^2^ = 15.18, *p* = 0.002; see Table 2). The interaction between *group* and *test-type* was a significant predictor of accuracy (beta = −0.85, *z* = −3.28, *p* = 0.001). The accuracy log-odds decreased for social items in the bvFTD group (OR = 0.43). In other words, after controlling for word frequency and overall semantic functioning: (a) SD and bvFTD participants had equal accuracy on non-social items, (b) SD participants were equally impaired on both social and non-social items, but (c) bvFTD participants were disproportionately *worse* for social items. As with the baseline model, both *semantic functioning* (beta = 1.22, *z* = 4.59, *p* <0.0001) and word *frequency* (beta = 0.33, *z* = 4.30, *p* < 0.0001) were significant predictors of accuracy in this model.

**Table 2.**
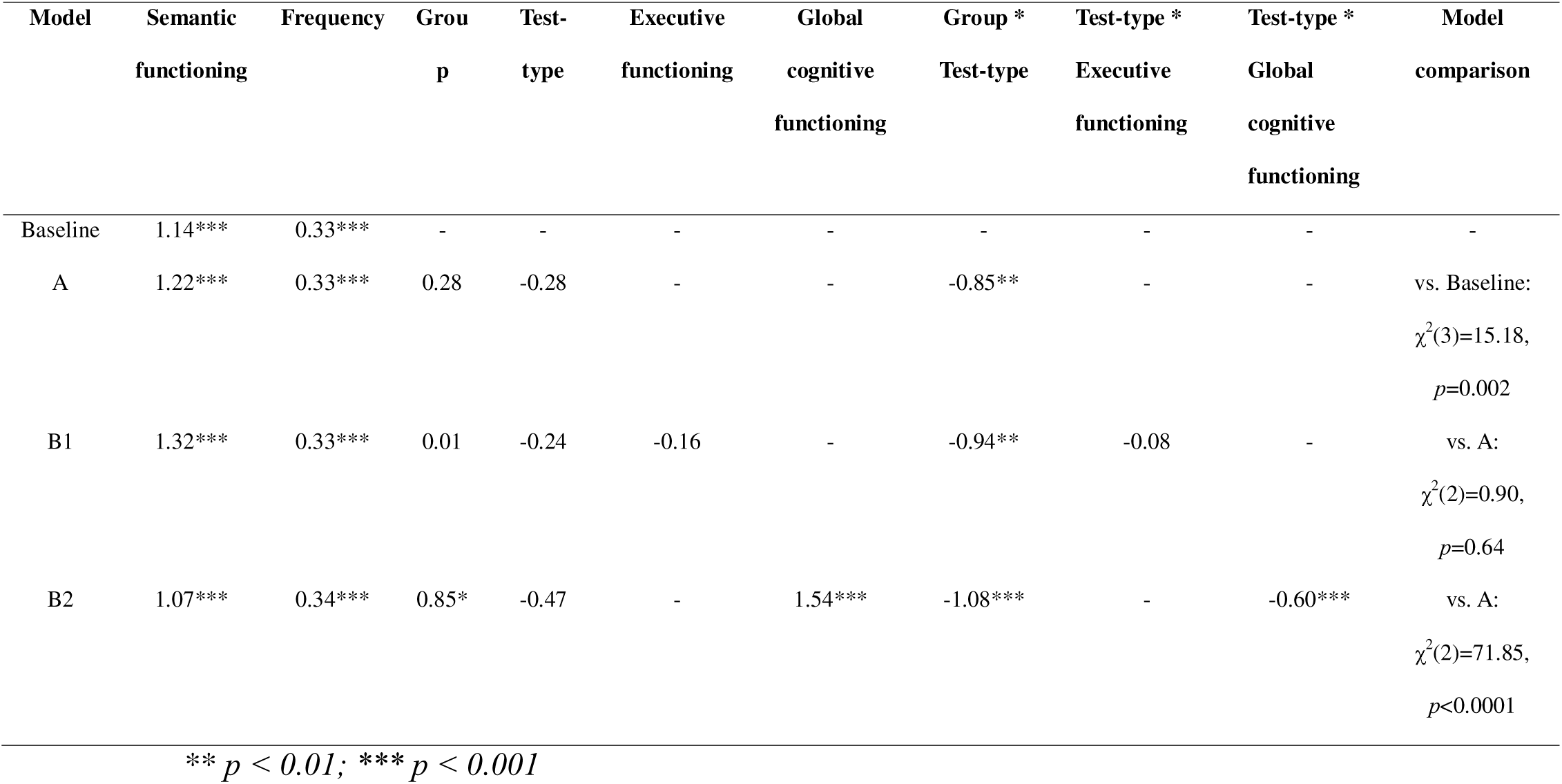
Beta coefficients for each predictor included in the linear mixed effects models.

| Model | Semantic<br>functioning | Frequency | Group<br>p | Test-type | Executive<br>functioning | Global<br>cognitive<br>functioning | Group *<br>Test-type | Test-type *<br>Executive<br>functioning | Test-type *<br>Global<br>cognitive<br>functioning | Model<br>comparison |
| --- | --- | --- | --- | --- | --- | --- | --- | --- | --- | --- |
| Baseline | 1.14*** | 0.33*** | - | - | - | - | - | - | - | - |
| A | 1.22*** | 0.33*** | 0.28 | -0.28 | - | - | -0.85** | - | - | vs. Baseline:<br>$\chi^2(3)=15.18$,<br>$p=0.002$ |
| B1 | 1.32*** | 0.33*** | 0.01 | -0.24 | -0.16 | - | -0.94** | -0.08 | - | vs. A:<br>$\chi^2(2)=0.90$,<br>$p=0.64$ |
| B2 | 1.07*** | 0.34*** | 0.85* | -0.47 | - | 1.54*** | -1.08*** | - | -0.60*** | vs. A:<br>$\chi^2(2)=71.85$,<br>$p<0.0001$ |

\*\* $p < 0.01$ ; \*\*\* $p < 0.001$

### Controlling for neuropsychological factors

A noted earlier, patients with SD and bvFTD differ in their neuropsychological profiles, potentially contributing to any observed group differences. For instance, bvFTD participants had increased levels of impaired executive function compared to SD, raising the possibility that the *group* by *test-type* interaction occurred because the social items place higher executive demands than the non-social items. If this is true, then the addition of *executive functioning* to the model would (a) contribute to prediction accuracy especially on social items and (b) eliminate the *group* by *test-type* interaction. We tested this possibility in Model B1:

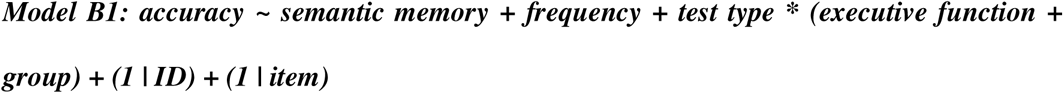

*Executive function* was not a significant predictor of accuracy (beta = −0.16, *z* = −0.60, *p* = 0.55), nor was the interaction between *executive functioning* and *test-type* (beta = −0.08, *z* = - 0.49, *p* = 0.63). The *group* by *test-type* interaction remained significant (beta = −0.94, z = - 2.89, p = 0.003). The inclusion of *executive functioning* did not improve model fit (χ^2^ = 0.90, *p* = 0.64) and so the term was excluded from the model.

Next, we considered whether the *group* by *test-type* interaction could be due to differences in *global cognitive functioning* between the two subtypes. Though there is no mean difference in overall cognitive functioning between the two groups, nevertheless social items might rely disproportionately on general cognitive functioning - in which case this factor should both (a) predict additional variation especially for social items and (b) eliminate the significant interaction of *group* by *test-type*. We tested this in Model B2:

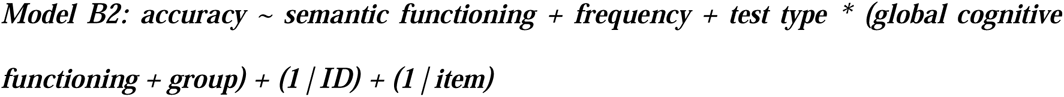

*Global cognitive functioning* was a significant predictor of accuracy log-odds (beta = 1.54, *z* = 10.56, *p* < 0.0001) indicating that that accuracy increased when cognitive functioning was better overall (OR = 4.68). The *global cognitive functioning* by *test-type* interaction was also significant, with a negative coefficient (beta = −0.60, *z* = −4.42, *p* < 0.0001), indicating that the benefit of better cognition overall was *reduced* for the social items. Critically, the *group* by *test-type* interaction not only remained after the inclusion of *global cognitive severity* but in fact got stronger (beta = −1.08, *z* = −3.91, *p* < 0.0001). A direct comparison between Model B2 and Model A showed that Model B2 had better fit (χ^2^ = 71.85, *p* < 0.0001) and so Model B2 was retained as the final model. Model statistics are shown in Table 2 and odds ratios of each predictor in the final model are displayed in Fig. 3.

**Fig. 3.**
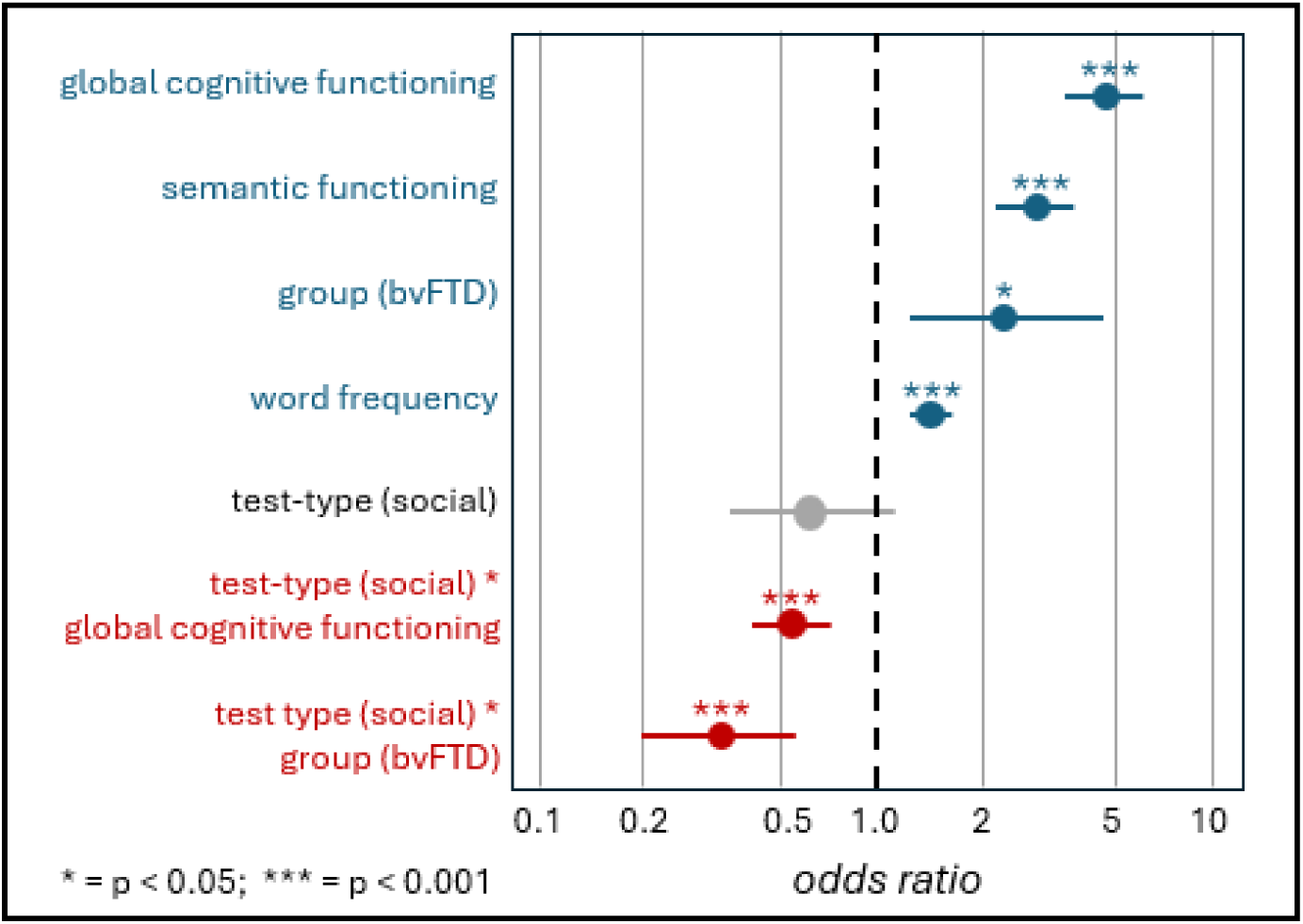
Odds ratios for predictors in the final binomial mixed-effects model predicting itemwise accuracy on the social-roles task. The reference condition is performance by patients with SD on non-social items. Better global cognitive functioning predicts higher accuracy generally, but the negative interaction with test-type (social) shows that this effect is reduced for social items. Better semantic functioning and higher word-frequencies predict higher accuracy, unsurprisingly. The null effect for test-type (social) indicates that performance on social items is not overall worse than on non-social items. Critically, however, the bvFTD group is marginally better than the SD group on the reference non-social items (positive group (bvFTD) effect), but significantly worse than the SD group on the social items (negative test-type (social) by group (bvFTD) interaction).

#### 3.2.3 Mixed-linear lesion-symptom correlation analysis

The significant interaction between test-type and diagnostic group in the behavioural analysis suggests that the two patient types differ on average in the robustness of social knowledge once confounding stimulus and neuropsychological factors are controlled. Yet the binary diagnosis measure in this analysis is a somewhat coarse, discrete shorthand for whatever underlying neuropathology is causing the knowledge impairment in each patient.

Indeed, the overlapping atrophy between SD and bvFTD groups (Fig. 1) indicates that the two diagnostic groups do not cleanly separate pathology in ATL vs. prefrontal regions. Ideally, we wish to see where cortical grey matter integrity predicts differential patterns of impairment on social versus non-social items, after removing effects of confounding factors, and regardless of the diagnostic category.

To this end we developed a mixed-linear-effects approach to lesion-symptom mapping. The logic is as follows: for every voxel location in the cortical mask, we fitted model B2 to predict participant accuracy on the social-roles item, but *replacing* the patient’s binary diagnostic grouping with a (continuous) measure of the patient’s grey matter intensity (*GMI*) at that voxel. If pathology at that voxel is associated with different degrees of impairment on social versus non-social items after accounting for other effects, then the *GMI* by *test-type* interaction should significantly improve model fit, compared to a reduced model that excludes the interaction. Thus, for every voxel in the cortical mask, we compared the fit of a full model:

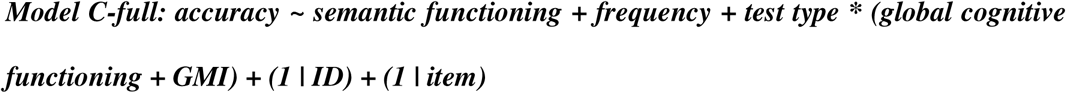

…to a reduced model that removed the test-type by GMI interaction:

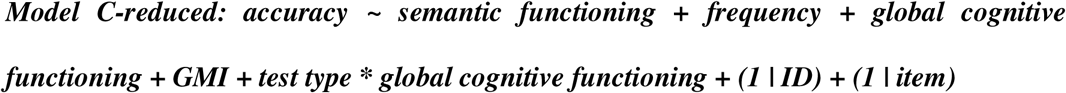

At each voxel we then stored the p-value associated with this contrast, indicating the probability that inclusion of the GMI by test-type interaction improves model fit over the reduced model - which in turn indicates that GMI at that location predicts differential accuracy outcomes for social versus non-social items, after accounting for all other effects.

We applied this analysis across all FTD participants with available structural MRI (SD = 18, bvFTD = 14), storing the contrast p-value at each voxel location. We then performed standard cluster-based controls for multiple comparisons to identify cortical regions where pathology consistently predicts different accuracy patterns on social and non-social items.

The results are shown in Fig. 4 and reported in Table 3. The analysis identified a broad region in the left insula and left lateral orbitofrontal cortex (Brodmann area 10), consistent with the OFC-spoke hypothesis. It further identified other regions often implicated in social cognition, including bilateral superior and middle frontal gyri and anterior cingulate cortex, as well as the supplementary motor area. Contradicting the social knowledge hypothesis, the mixed-linear analysis did not implicate any ATL subregion - despite the strong correlation between ATL pathology and general semantic impairment shown in Fig. 2.

**Fig. 4.**
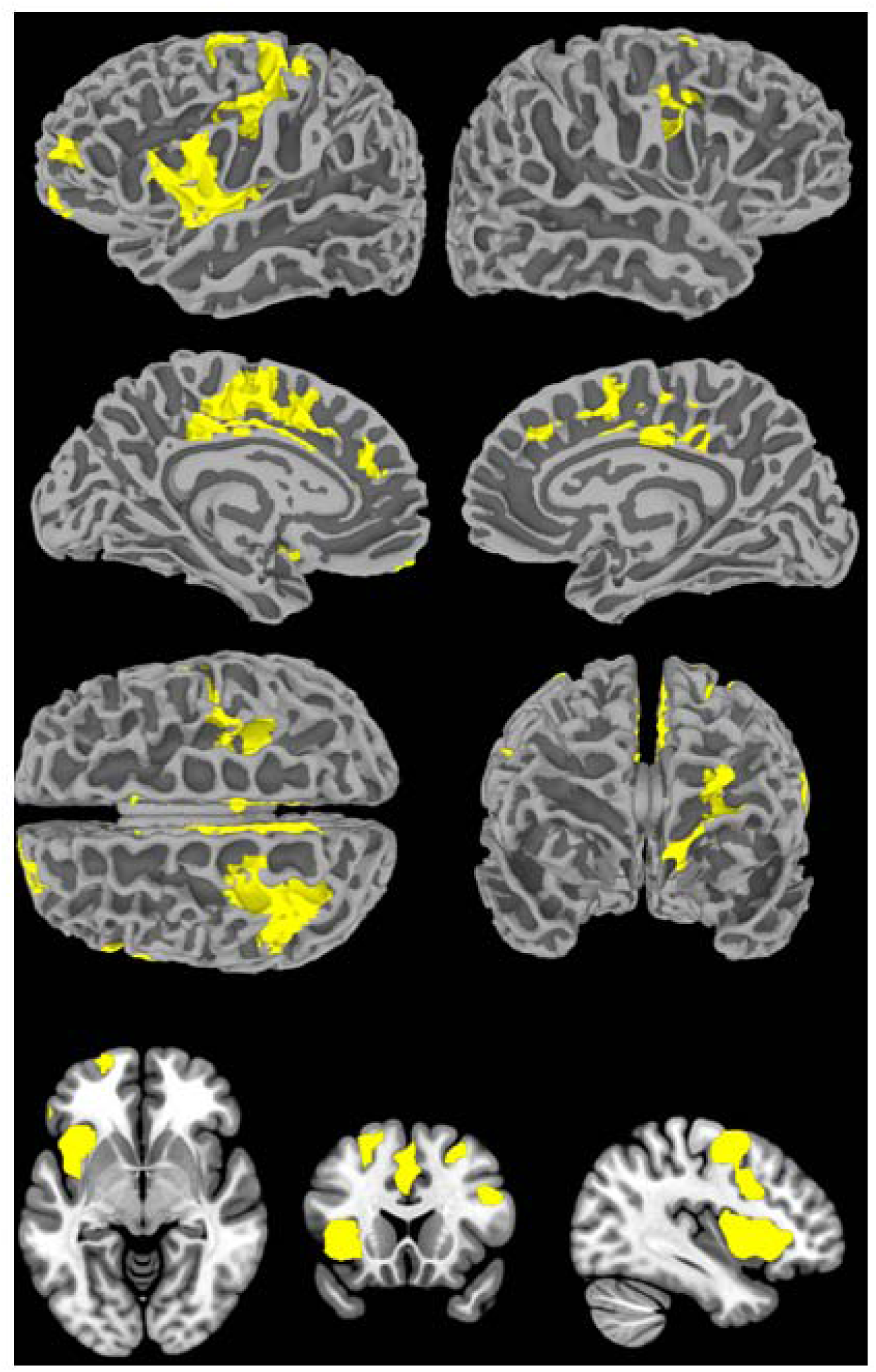
Cortical regions revealed by the mixed-linear lesion-symptom mapping analysis. Coloured areas indicate regions where grey matter intensity interacts with stimulus type in the mixed-effects model, predicting different levels of impairment on social versus non-social test items after other effects have been regressed out. The top three rows show results projected to a standard cortical surface model while the bottom row shows results on slices through a standard brain volume. The map identifies left OFC, left insula, bilateral superior frontal, and cingulate regions. Note that all areas differ from the maps in Fig. 2, showing regions where cortical atrophy correlates with overall cognitive functioning and with semantic functioning. Also note that the analysis does not identify any sub-regions of ATL as being differentially important for social concepts.

**Table 3.** Brain regions associated with a disproportionate social conceptual impairment.

| Regions | Hemisphere | Number of voxels | Peak MNI co-ordinate |  |  | Peak structure | z-score |
| --- | --- | --- | --- | --- | --- | --- | --- |
|  |  |  | x | y | z |  |  |
| Precentral gyrus, middle frontal gyrus, superior frontal gyrus, inferior frontal gyrus, postcentral gyrus | Left | 627 | -30 | 0 | 51 | Middle frontal gyrus | -4.06 |
| Middle cingulate gyrus, supplementary motor area, anterior cingulate gyrus, superior frontal gyrus, posterior cingulate gyrus | Bilateral | 616 | -9 | -12 | 57 | Supplementary motor area | -3.73 |
| Insula, inferior frontal gyrus, Rolandic operculum, olfactory cortex, superior temporal gyrus | Left | 647 | -39 | 18 | -3 | Insula | -3.64 |
| Middle frontal gyrus, superior frontal gyrus | Right | 96 | 27 | 15 | 45 | Middle frontal gyrus | -3.32 |
| Inferior frontal gyrus, precentral gyrus, middle frontal gyrus | Right | 111 | 36 | 9 | 36 | Inferior frontal gyrus | -2.96 |
| Superior frontal gyrus, middle frontal gyrus | Left | 77 | -24 | 66 | -6 | Superior frontal gyrus, medial orbital | -3.12 |
| Superior frontal gyrus, anterior cingulate gyrus, middle cingulate gyrus | Bilateral | 61 | 3 | 42 | 27 | Anterior cingulate cortex | -3.01 |

## 4. Discussion

The main outcome of this study is to have resolved the neural basis of social concepts by testing three distinct hypotheses: (1) the *social-knowledge* view, which proposes that social knowledge relies preferentially on subregions of the ATLs, (2) the *ATL-hub* view, which proposes that social and non-social concepts alike are supported by a domain-general semantic hub within the ATLs, and (3) the *ATL-hub + social-spokes* view, which proposes that ventral prefrontal cortices and insula contribute more to social than non-social knowledge by virtue of their connection to/involvement in computations important to social concepts such as motivation, affect, and reward. Prior work does not clearly adjudicate these different views because assessments of social and non-social knowledge often differ along multiple factors potentially impacting performance; because the distinction between social and non-social knowledge is somewhat ill-defined and variable across studies; and because different patient populations can exhibit quite different varieties of cognitive dysfunction.

Our aim in this paper was to test the three hypotheses via a new assessment of social and non-social knowledge in FTD that clearly distinguishes these domains, using analysis techniques that control or regress out many potential confounding factors. The results offer strong support for the ATL hub + social-spokes hypothesis, in two respects. First, the mixed-linear analysis of patient behaviour showed that, after accounting for both itemwise factors (e.g. word frequency) and neuropsychological factors (general cognitive functioning and semantic functioning), patients with SD show equivalent degrees of impairment on social and non-social items whereas those with bvFTD show significantly better performance on non-social items and significantly worse performance on social items. If all concepts are supported solely by a domain-general ATL hub, no difference should arise between social and non-social items for either group after other effects are regressed out; and if ATLs house a region dedicated to social knowledge, then such knowledge should be worse in SD (where ATL pathology is extensive) than in bvFTD (where it tends to be less pervasive; see Fig. 1). Instead, we observed the reverse interaction, with worse performance on social concepts observed for the bvFTD but not the SD group, consistent with the ATL-hub + social-spoke view.

Second, the mixed-linear lesion-symptom mapping analysis found that different patterns of impairment on social versus non-social items were associated with greater pathology in OFC and insula regions, together with other cortical areas often implicated in social knowledge, but *not* with any subregion of the ATL. This too accords with the ATL-hub + social-spoke view and contradicts both alternative views. If social concepts were solely supported by a domain-general hub, we would expect no brain regions to consistently show this pattern; and if social concepts relied on a dedicated ATL subregion, we would expect such a region to appear in the map.

In the following sections, we discuss these two key findings and how they relate to a broader understanding of conceptual representation in the brain.

### 4.1 The anterior temporal lobe and social concepts

#### 4.1.1 A shared bilateral ATL hub for social and non-social concepts

People with SD experience a progressive deterioration of conceptual knowledge, for all types of concepts and across all receptive and expressive modalities, following bilateral ATL atrophy (Bozeat et al., 2000; Hodges et al., 1992; Mummery et al., 2000; Patterson et al., 2007). The current work provides further evidence that the impairment also encompasses social concepts. This finding aligns with our previous study which investigated of a wide range of social concepts in a large FTD cohort, and found that performance on social and non-social tasks was highly correlated and associated with grey matter volume in the bilateral ATL (Rouse, Halai, et al., 2024).

Several additional sources of evidence further support the view that the ATL hub supports semantic knowledge in a broad, domain-general way. First, people with unilateral left or right ATL resection to treat temporal lobe epilepsy display a mild generalised semantic deficit that occurs for all types of concept, including social concepts (Lambon Ralph et al., 2012; Rice, Caswell, et al., 2018; Rouse, Halai, et al., 2024; Rouse, Ramanan, et al., 2024). Second, fMRI studies in healthy participants have found that social and non-social semantic tasks are associated with overlapping BOLD activation centred on the bilateral ventrolateral ATL (Binney et al., 2016; Rice, Hoffman, et al., 2018). Finally, transcranial magnetic stimulation to the ATL disrupts performance on both social and non-social semantic tasks (Pobric et al., 2016).

Some work proposes that left and right ATLs contribute differently to conceptual representation with, for instance, the right ATL supporting social concepts and the left supporting verbal semantics (Younes et al., 2022; Zahn et al., 2009). This hypothesis is based largely on the consistent clinical observation that the minority of SD patients who initially have asymmetric right > left atrophy in ATL typically present with face recognition problems and behavioural changes (Ding et al., 2020; Evans et al., 1995; Gorno-Tempini et al., 2004). The current work found that both social and non-social conceptual knowledge were associated with *bilateral* ATL volume (Fig. 2), consistent with previous studies (Rijpma et al., 2023; Rouse, Halai, et al., 2024). We found no evidence that right ATL atrophy led to a disproportionate impairment for social concepts (Fig. 4). Instead, the selective social impairment was related to atrophy in prefrontal regions and the insula across the entire FTD cohort. Thus, a different explanation of the relationship between R > L ATL atrophy and behavioural change is that this reflects correlated atrophy in insula and orbitofrontal regions (as suggested in previous studies) (Ding et al., 2020; Rouse, Binney, et al., 2024, 2024; Rouse et al., 2025). Indeed, multiple previous studies have shown that R > L presenting patients typically have more atrophy overall across the ATLs, insula and multiple prefrontal regions (Ding et al., 2020; Rohrer et al., 2008; Rouse, Halai, et al., 2024; Seeley et al., 2005). Finally, unilateral resection of the right ATL for temporal lobe epilepsy does not generate a selective social concept deficit. Instead, left *or* right ATL resection yields a subtle semantic impairment that occurs for all aspects of conceptual knowledge, including social concepts (Lambon Ralph et al., 2012; Rice, Caswell, et al., 2018; Rouse, Halai, et al., 2024; Rouse, Ramanan, et al., 2024). Consistent with this view, a large scale meta-analysis of fMRI studies found no evidence of ATL laterality differences for social and non-social stimuli in healthy participants (Rice, Lambon Ralph, et al., 2015), and TMS to the left or right ATL disrupts both social and non-social semantic processing (Pobric et al., 2016). From a functional perspective, computational modelling has shown that a bilaterally-implemented hub becomes much more resilient to unilateral damage (Schapiro et al., 2013), suggesting one reason why the system may have evolved this way (Ding et al., 2020; Jung et al., 2021; Rouse, Ramanan, et al., 2024).

### 4.2 Beyond the ATL

The above considerations establish that ATLs support knowledge for both social and non-social domains. The current work further suggests that the ATLs interacts with “social spokes” in insula, OFC, and other areas, that support social conceptual knowledge somewhat more than non-social knowledge.

The idea that graded functional specialisation arises within the hub-and-spokes semantic network mainly stems from patterns of modality-specific responding observed across the superior-to-ventral axis of the ATLs. Thus, while the full ATLs contribute to conceptual knowledge across modalities, more superior aspects that are tightly interconnected to perisylvian language areas (a “language spoke”) respond more strongly to verbal than to visual semantic tasks, whilst the reverse is true for ventromedial ATL regions tightly interconnected to the ventral visual stream (a visual object “spoke”). The anterior extent of the fusiform gyrus, lying between these regions, responds equally in visual and verbal tasks, and forms the nexus of pathology causing cross-modal impairment in SD (Lambon Ralph et al., 2017; Patterson et al., 2007). From these ideas, it has been proposed that the ATLs adopt graded functional specialisation based on the extent of their connectivity to other representational systems (Rice, Hoffman, et al., 2015). We propose that the insula, OFC, and other areas shown in Fig. 4 may constitute a social “spoke” that contributes to semantic knowledge in a graded manner, with somewhat more importance for social than non-social concepts.

The infero-frontal and insula regions that appear differentially important for social versus non-social knowledge in the current work certainly constitute an anatomical “spoke” connected to the ATL: insula lies immediately beneath superior ATL, while OFC is directly connected to its superior and lateral aspects via the uncinate fasciculus (Cloutman et al., 2012; Papinutto et al., 2016). The central remaining question is why these regions might be disproportionately important for social knowledge - particularly the form of social knowledge assessed by the social roles test, namely the meanings of words referring to classes of people. What makes the regions identified in Fig. 4 “social”?

Presumably the answer has to do with the nature of the representations or computations implemented by these areas, which are more likely to be engaged when participants process information about people versus objects. Thus, we can ask, what do we know about the corresponding representations and processes, and how might these be engaged by social vs. non-social processes? This section briefly reviews current understanding of the functions of the key areas and considers whether and how they may be preferentially important for social knowledge. We note that these ideas are somewhat speculative and, we hope, lay foundations for future work.

*The insula and interoceptive input.* The finding that insula pathology correlates with differential performance on social vs. non-social items mirrors a previous study where a selective correlation was found between insula volume and social-semantic performance in FTD (Rijpma et al., 2023). The insula is a key node in the *salience* network, a large-scale distributed network of brain areas systematically affected in bvFTD, where it is thought to be critical for alerting individuals to personally salient signals to guide socially appropriate behaviour (Seeley et al., 2009).

How can this finding be integrated within the hub-and-spokes model? The insula is understood to play an important role in interoceptive processing, that is, the ability to integrate internally-generated signals about bodily states (e.g. hunger/thirst, emotional states; see Critchley et al. 2004). Thus the insula may integrate interoceptive signals to generate global feeling states (Ibañez & Manes, 2012; Seeley et al., 2012). The insula may then serve as a “social spoke” by providing the ATL with interoceptive information of particular importance to social behaviours. For instance, concepts such as *grandmother* and *baby* may evoke greater emotional valence than non-social concepts like *shed* or *piano*, detected by the insula as part of its interoceptive role and communicated to the ATL. In support of this idea, the ‘interoceptive strength’ of a concept (i.e., degree to which a concept is experienced through sensations in the body) is higher for emotion concepts than for neutral (Connell et al., 2018), Moreover, TMS to the anterior insula in healthy participants disrupts performance on a semantic task using emotion and social concepts, but not non-social concepts (Mancano & Papagno, 2026).

*The OFC: Computation of value or nexus of social control?* The OFC is often understood to represent/compute the *subjective value* of stimuli or behaviours (O’Doherty et al., 2001; Padoa-Schioppa & Assad, 2006) - therefore the region may serve as a social spoke to the extent that social items are more likely than non-social items to evoke significant positive or negative reward value. Here again the emotional valence associated with reward may be a key ‘ingredient’ for concepts relating to knowledge about people (Warriner et al., 2013). In support of this idea, a recent study found that words with high ratings of ‘socialness’ had higher ratings of valence extremity, indicating that social concepts rely more on affective information compared to general non-social concepts (Diveica et al., 2023).

However, there are several important differences between valence and other sources of information that must be taken into consideration whilst conceptualising the orbitofrontal cortex as a ‘social spoke’. Unlike other sources of modality-specific information, value is not a sensory property of a concept that can be directly experienced. Rather, the value of any object is highly dependent on its meaning and context: e.g., if one is hungry then the value of a small spherical object will be higher if it is an apple as opposed to a tennis ball. This implies that value computation requires a detailed activation of the concept. Accordingly, it would then follow that in addition to the ATL receiving OFC-based value information, the ATL-based semantic system provides a key input to orbitofrontal-based value computations. Second, value needs to be rapidly and flexibly updated based on changing contexts (O’Doherty et al., 2000). For example, the subjective value of a strawberry will decline if eaten to satiety and will also vary based on its current state (if a strawberry is ripe, it will have a higher value than if it is mouldy). Extending this proposal to social concepts, tears can either be negatively valenced (e.g., at a funeral) or positively valenced (e.g., at a comedy night) depending entirely on the situational context. It should be noted that value is not a property exclusive to social concepts (going back to the previous example, value is also associated with food – another known type of behaviour change common in FTD) (Rascovsky et al., 2011), but is, nevertheless, centrally relevant to social concepts which we typically ‘tag’ with emotional valence.

Alternatively, the OFC may play a key role in the *control* of social conceptual knowledge, guiding socially appropriate behaviour in response to changing reward contingencies. S*ocial control* refers to the selection, evaluation, value-based decision making and inhibition processes that interact with semantic representations for the generation, implementation or inhibition of adaptive social behaviours across changing social settings (Rouse, Halai, et al., 2024). Alongside the anterior cingulate cortex, the OFC is thought to have an important role in representing the value of objects and actions, which interacts with ATL-mediated social conceptual knowledge to shape social behaviour.

### 4.3 Semantic control network

In addition to the insula and OFC, differential impairment to social and non-social items was predicted by atrophy in the lateral prefrontal cortex, anterior cingulate cortex and supplementary motor area. These brain regions form part of the neural network that supports *semantic control* - the specifically-targeted executive processes that manipulate semantic representations to shape context-appropriate behaviours (Jackson, 2021; Lambon Ralph et al., 2017). Stronger input from the semantic control network is required when task demands require the retrieval of weakly encoded information, suppression of over-learned responses or emphasis of uncharacteristic features (Jefferies & Lambon Ralph, 2006), and also for different types of concept that have some of these same weaknesses (e.g., comprehension of abstract words requires higher semantic control than comprehension of concrete words) (Hoffman, 2016; Hoffman, Lambon Ralph, et al., 2013). It is possible is that the social concepts task placed higher demands on semantic control than the non-social task comparator task. For example, unlike the manmade items where the picture directly identifies the concept (e.g., shed), the social roles are only ever implied by the picture (e.g., the picture of an older woman could point to many different meanings and senses including, but not limited to, “grandma”). It is perhaps important to note that at least some parts of the semantic control network are distinct from those associated with general “multi-demand” executive areas of the brain (Hodgson et al., 2024; Humphreys & Lambon Ralph, 2017). Thus, although we controlled for executive function, we did not include any specific tests of semantic control which means we are unable to rule out the degree to which the social impairment in bvFTD partially reflects a semantic control problem. Further research is required to determine whether increased semantic control demands are a feature common to social concepts more broadly or are specific to the task employed here.

### 4.4 Limitations and future directions

The current study used only one type of social stimuli – words describing social roles. However, social concepts span a very diverse range of concrete-to-abstract concepts, including people but also including social behaviours and emotions. From this study, is it not possible to ascertain the degree to which the brain regions associated with social conceptual deficits are equally critical for other types of social concept. Consequently, future work could apply the same linear mixed effects models to other exemplars of social concepts, including abstract words that describe social behaviours (e.g. *honesty* and *greed)* (Binney et al., 2016).

The overlap between the semantic control network and the brain regions associated with the social conceptual deficit opens up two intriguing avenues of neuropsychological enquiry. First, future research should investigate whether the known executive function impairment in bvFTD extends to semantic control, and the extent to which the social semantic impairments in this patient group can be explained by increased semantic control demands. Second, future work should explore whether people who have known semantic control deficits due to post-stroke aphasia have concurrent impairments for social concepts.

The patient group studied here have a combination of ATL and prefrontal-insula damage. Indeed, the mixed effects modelling includes metrics associated with both factors; and the hub-and-spoke implied important conjoint action in all concepts. Thus, we cannot tell if the additional effect for social concepts requires/is conditional of the underlying ATL damage – or if it is independent of it. This would require future comparative studies in which patients with selective insula/OFC but without ATL damage, are directly compared with the bvFTD and SD cases (an approach taken previously for comparing SD bilateral ATL patients with unilateral ATL cases following surgical resection) (Rouse, Halai, et al., 2024; Rouse, Ramanan, et al., 2024).

## 5. Conclusion

Our results demonstrate that the bilateral ATL is critical for all aspects of semantic memory, including social concepts. An additional disproportionate impairment for social concepts is associated with atrophy in brain regions associated with bvFTD, including the insula and prefrontal cortex. Our findings converge on the hub-and-spokes model of semantic representation, in which the bilateral ATLs underpin a transmodal semantic hub, and secondary differences emerge between social and non-social concepts based on their differing reliance on either modality-specific spokes or control processes.

## Data Availability

Owing to the limits of the ethics approval for these patient studies, the raw data cannot be shared openly. Requests for anonymised data can be addressed to the senior authors and may require a data transfer agreement.

## Credit author statement

Matthew Rouse: Conceptualization, Data curation, Formal analysis, Investigation, Methodology, Project administration, Software, Visualization, Writing – original draft; Peter Garrard: Resources, Writing – review & editing; James Rowe: Resources, Writing – review & editing; Matthew Lambon Ralph: Conceptualization, Methodology, Supervision, Writing – review & editing; Timothy Rogers: Conceptualization, Data curation, Methodology, Software, Supervision, Writing – review & editing.

## Acknowledgements

We thank the patients and their families or caregivers for giving up the time to take part in the study. We also thank Prof Masud Husain, Dr Sian Thompson and Dr Sofia Toniolo for their help with recruitment.

