## Supplementary Material for "Social concepts rely on a domain-general anterior-temporal hub and social “spokes” in ventral prefrontal cortex and insula"

Supplementary Table 1. Voxel-based morphometry results

| **Regions** | **Hemisphere** | **Number of voxels** | **Peak MNI co-ordinate** | | | **Peak Structure** | ***t*-value** |
|---|---|---|---|---|---|---|---|
| | | | ***x*** | ***y*** | ***z*** | | |
| *SD < Controls* | | | | | | | |
| | Bilateral | 1,555,880 | -29 | -6 | -41 | Left inferior temporal gyrus | 22.48 |
| | Right | 2,539 | 29 | -69 | -48 | Lobule VIII of cerebellar hemisphere | 4.91 |
| *bvFTD < Controls* | | | | | | | |
| | Bilateral | 1,283,950 | 2 | 36 | -15 | Gyrus rectus | 8.56 |

bvFTD = behavioural-variant frontotemporal dementia, MNI = Montreal Neurological Institute, SD = semantic dementia

Supplementary Table 2. Mean scores on each background neuropsychology task and composite indices

| | **SD** | **bvFTD** | **Control** | **Group difference** | **Post-hoc** |
|---|---|---|---|---|---|
| N | 21 | 24 | 19 | - | - |
| ACE-R Total (100) | 47.5 (22.0) | 63.6 (19.2) | 96.8 (2.3) | **F(2,61)=42.1, p<0.0001** | bvFTD, SD < C, SD < bvFTD |
| Mini Mental State Examination (30) | 20.0 (8.1) | 22.5 (5.5) | 29.8 (0.4) | **H(2)=35.3, p<0.0001** | SD, bvFTD < C |
| ACE-R Attention (18) | 13.1 (5.2) | 14.3 (3.8) | 17.9 (0.2) | **H(2)=27.7, p <0.0001** | SD, bvFTD < C |
| ACE-R Memory (26) | 8.5 (6.4) | 14.2 (7.4) | 24.5 (2.0) | **H(2)=36.0, p<0.0001** | SD, bvFTD < C, SD < bvFTD |
| ACE-R Fluency (14) | 4.7 (3.4) | 4.4 (3.3) | 13.2 (1.2) | **H(2)=40.0, p<0.0001** | SD, bvFTD < C |
| ACE-R Language (26) | 9.2 (5.1) | 18.9 (6.9) | 25.7 (0.5) | **H(2)=44.2, p<0.0001** | SD, bvFTD < C; SD < bvFTD |
| ACE-R Visuospatial (16) | 12.0 (4.6) | 11.9 (3.5) | 15.6 (0.8) | **H(2)=64, p<0.0001** | SD, bvFTD < C |
| Cambridge Naming (32) | 13.6 (9.5) | 28.5 (6.9) | 31.9 (0.2) | **H(2)=45.3, p<0.0001** | SD < bvFTD, C |
| Boston Naming (30) | 7.1 (5.4) | 22.6 (7.8) | 29.7 (0.5) | **H(2)=47.0, p<0.0001** | SD < bvFTD, C; bvFTD < C |
| Camel and cactus test (32) | 15.7 (5.1) | 21.8 (7.4) | 30.7 (1.1) | **H(2)=34.3, p<0.0001** | SD < bvFTD, C; bvFTD < C |
| Synonym judgement (48) | 35.9 (7.5) | 39.0 (7.9) | 47.8 (0.4) | **H(2)=37.60, p<0.0001** | SD, bvFTD < C |
| Brixton (10) | 4.8 (2.8) | 2.9 (2.0) | 6.4 (2.0) | **H(2)=19.3, p<0.0001** | bvFTD < C, SD |
| Raven's (12) | 8.3 (3.4) | 5.0 (2.7) | 10.5 (1.5) | **H(2)=27.67, p<0.0001** | bvFTD < SD; bvFTD, SD < C |
| Face-name matching (44) | 16.4 (7.9) | 28.6 (11.1) | 38.9 (3.4) | **H(2)=29.29, p<0.0001** | SD < bvFTD; bvFTD, SD < C |
| Face-profession matching (44) | 20.0 (10.0) | 27.7 (11.4) | 40.3 (3.7) | **H(2)=25.89, p<0.0001** | SD < bvFTD; bvFTD, SD < C |
| Landmark-name matching (42) | 16.1 (6.3) | 24.3 (9.0) | 38.5 (1.8) | **W(2, 28.8)=130.15, p<0.0001** | SD < bvFTD; bvFTD, SD < C |
| Famous face matching (22) | 18.0 (2.1) | 18.6 (2.8) | 21.2 (0.8) | **H(2)=22.56, p<0.0001** | bvFTD, SD < C |
| Unfamiliar face matching (22) | 18.1 (2.8) | 17.1 (3.1) | 20.3 (1.4) | **H(2)=13.49, p = 0.001** | bvFTD, SD < C |
| Social abstract synonym judgement (36) | 25.1 (5.8) | 26.4 (5.3) | 33.9 (1.3) | **F(2,55)=20.20, p<0.0001** | bvFTD, SD < C |
| Non-social abstract synonym judgement (36) | 25.5 (6.7) | 28.1 (6.1) | 35.6 (0.6) | **H(2)=32.7, p<0.0001** | bvFTD, SD < C |
| Basic emotion matching (19) | 11.0 (3.5) | 11.8 (3.2) | 16.3 (1.5) | **F(2,53)=18.8, p<0.0001** | bvFTD, SD < C |
| Complex emotion matching (23) | 12.2 (5.0) | 12.9 (4.9) | 18.5 (1.9) | **F(2,52)=12.9, p<0.0001** | bvFTD, SD < C |
| Social Norms Questionnaire (22) | 15.5 (2.7) | 15.4 (4.0) | 20.0 (1.2) | **H(2)=22.1, p<0.0001** | bvFTD, SD < C |
| TASIT-Sarcasm (20) | 8.1 (5.1) | 9.9 (5.5) | 18.7 (1.9) | **H(2)=29.8, p<0.0001** | bvFTD, SD.< C |
| Semantic functioning | -0.66 (0.59) | 0.63 (0.94) | - | **t(39.2) = 5.46, p <0.0001** | - |
| Global cognitive functioning | 0.25 (1.01) | 0 (0.83) | - | t(43) = 0.92, p = 0.36 | - |
| Executive functioning | 0.53 (1.0) | -0.44 (0.83) | - | **t(43) = 3.57, p = 0.0009** | - |

Means and standard deviations reported for each group. Significant P-values are bolded.
ACE-R, Addenbrooke's Cognitive Examination-Revised; bvFTD, behavioural-variant frontotemporal dementia; C, control; FTD, frontotemporal dementia; SD, semantic dementia; TASIT, The Awareness of Social Inference Test.

Supplementary Table 3. Regions of grey matter associated with neuropsychological components

| Regions | Hemisphere | Number of voxels | Peak MNI co-ordinate | | | Peak Structure | t-value |
|---|---|---|---|---|---|---|---|
| | | | x | y | z | | |
| *Global cognitive functioning* | | | | | | | |
| Middle frontal gyrus, superior frontal gyrus, inferior frontal gyrus, precentral gyrus | Right | 3,638 | 30 | 27 | 56 | Superior frontal gyrus | 6.42 |
| Middle frontal gyrus, superior frontal gyrus, inferior frontal gyrus, supplementary motor area, precentral gyrus, middle cingulate cortex, caudate nucleus, inferior parietal lobule, Rolandic operculum, putamen, postcentral gyrus, insula, anterior orbitofrontal cortex, medial orbitofrontal cortex, superior anterior cingulate cortex, superior orbitofrontal cortex, lateral orbitofrontal cortex, medial orbitofrontal cortex, Heschl's gyrus, superior temporal gyrus, superior temporal pole, gyrus rectus, supramarginal gyrus, pregenual anterior cingulate cortex, thalamus, nucleus accumbens, globus pallidus, olfactory cortex | Left | 36,196 | -41 | 5 | 36 | Precentral gyrus | 5.96 |
| *Semantic functioning* | | | | | | | |
| Inferior temporal gyrus, fusiform gyrus, middle temporal gyrus, superior temporal pole, middle temporal pole, parahippocampal gyrus, superior temporal gyrus, hippocampus, amygdala, cerebellum lobule IV-V, insula, cerebellum lobule VI, inferior frontal gyrus, cerebellum lobule III, lingual gyrus, olfactory cortex, cerebellum crus I | Right | 23,291 | 44 | 26 | -32 | Superior temporal pole | 7.12 |

| | | | | | | | |
|---|---|---|---|---|---|---|---|
| Inferior temporal gyrus, middle temporal gyrus, fusiform gyrus, superior temporal pole, middle temporal pole, parahippocampal gyrus, superior temporal gyrus, insula, hippocampus, amygdala, cerebellum lobule VI, cerebellum lobule IV-V, olfactory cortex, putamen, inferior frontal gyrus, cerebellum lobule III, lingual gyrus, Rolandic operculum, globus pallidus, cerebellum crus I, Heschl's gyrus | Left | 23,484 | -44 | 5 | -15 | Superior temporal pole | 6.20 |
